# Pilot: Salivary Lactoferrin as a Biomarker of Alzheimer’s Disease

**DOI:** 10.1101/2024.11.22.24317405

**Authors:** BL Hammerschlag, B Butts, KDS Likos, DD Verble, N Nimmagadda, R Virani, S Ramanathan, W. Wharton

## Abstract

**Background:** Alzheimer’s disease (AD) research has focused on developing accessible biomarkers that accurately detect disease pathology and progression before symptoms present. Lactoferrin (Lf) is an iron-binding antimicrobial glycoprotein found in all biological fluids, and its concentration in saliva has been correlated with AD symptoms. This pilot project aimed to determine whether salivary lactoferrin (sLF) has potential as a biomarker for AD.

**Methods:** Participants were middle to older-aged non-Hispanic white (NHW) and Black Americans (BA) at risk for AD due to parental history. We collected saliva samples after an 8-hour fast and administered a cognitive battery assessing executive function, memory, visuospatial ability, attention, and verbal fluency. We examined the relationship between sLF and cognitive performance and evaluated protein concentration across races.

**Results:** Seventeen middle-to-older-aged (age = 60.29 ± 9.7 years) BA and NHWs were enrolled. After controlling for age, sex, race, and years of education, we found a significant r between sLF and Digit Span Memory Test (DSMT) scores (*P* = 0.013) and a modest correlation with Mental Rotation Test scores (*P* = 0.194). We found no difference in average concentration across races.

**Conclusions:** Memory concerns and a worsening in visuospatial ability are early signs of cognitive decline in AD patients, and this pilot suggests a correlation of these symptoms with sLF. Bigger-scale longitudinal studies to examine the relationship between sLF and established AD biomarkers in diverse populations are needed to assess its clinical usefulness as an early biomarker for AD.

## Introduction

Alzheimer’s Disease (AD) is the 7th leading cause of death in the United States (Center for Disease Control, 2022). Assessing risk and developing preventive mechanisms for AD is a top priority for healthcare research in the United States. Neuropathological changes linked to AD emerge many years before cognitive symptoms emerge (Dubois, 2016). Therefore, the presymptomatic disease stage provides a crucial window of opportunity both to understand the mechanisms through which AD evolves from pathogenesis to clinical symptoms and to potentially treat a patient before symptom onset and progression.

Imaging technologies such as PET and MRI and biomarkers in cerebrospinal fluid (CSF) and plasma, specifically amyloid–β (Aβ42) and phosphorylated tau (*p*-tau), are used to diagnose AD and guide clinical decisions throughout disease progression (Jack, 2024). However, imaging equipment, blood draws and lumbar punctures require trained staff, timely processing, and expensive equipment, with participants often having to travel to institutions (Ashton, 2021).

Saliva as a biofluid is a valuable alternative. Patients and research participants may favor it because it is simple and non-invasive; clinics and research institutions may favor it because collection can be performed by the participant without the need of trained clinical staff (Wan-Chen, 2023). Saliva concentrations of neurodegenerative disease biomarkers such as total tau (t-tau) and neurofilament light chain protein (NfL) are not disease specific for AD and do not correlate to plasma AD biomarkers (Ashton, 2018)(Gleerup, 2021). Salivary Lactoferrin, however, poses promising evidence.

Lactoferrin (Lf) is an iron-binding antimicrobial glycoprotein that can be found in most biological fluids, including all exocrine secretions in mammals (Bermejo-Pareja, 2020). It was first isolated from bovine milk in 1939 and later described to modulate immune responses with its anti-inflammatory properties (Ashton, 2019)(Actor, 2009). More recently, it was described as a key nutrient for neurodevelopment and cognitive function during early periods of rapid brain growth (Chen et al., 2015).

Lactoferrin may indirectly lessen AD risk by reducing neuroinflammation and protection from microbial infection. Lf is highly expressed in cortical tissue affected by AD pathology, specifically in senile plaques composed of aggregated β-amyloid (Aβ), extra- and intracellular neurofibrillary tangles, and glial cells of AD patients (Kawamata, 1993). Studies show that systemic viral and bacterial infections are significant risk factors for AD, and our group and others have reported increased neuroinflammation in patients with MCI/AD and individuals at high risk for AD (Wharton, 2019)(Leng, 2021)(Itzhaki, 2016). Lactoferrin, a known antimicrobial and anti-inflammatory protein, may exert neuroprotection and thus low levels or decreases in Lf may serve as an indicator of AD risk (Welling, 2015)(Rousseau, 2013)(Gleerup, 2019).

Animal studies show that Lf can ameliorate mitochondrial dysfunction and lower levels of known markers of neuroinflammation (Park et al, 2013)(Mayeur et al., 2016). Moreover, Lf administration significantly improved spatial cognition in old mice (Zheng et al., 2020) and reduced amyloid deposition and Aβ40 and Aβ42 in CSF (Guo et al., 2017)(Abdelhamid, 2020). In saliva, Antequera et al. reported a significant early and robust decrease of sLf in six- and twelve-month-old APP/PS1 mice that was not seen in their age-matched wild type counterparts (WT) (Antequera, 2021). Importantly, they did not see a decrease in total protein concentration in saliva.

Three clinical studies have shown a relationship between low sLF and cognitive decline in AD patients and healthy individuals at risk for AD. In an observational study among 116 patients with a diagnosis of MCI or AD, authors reported that sLf correlates with Mini-Mental State Examination (MMSE) scores and CSF biomarkers Aβ42 and total tau, and could differentiate healthy controls from participants with an MCI/AD diagnostic (Carro, 2017). Authors also described excellent performance differentiating MCI/AD versus frontotemporal dementia (Gonzalez-Sanchez, 2020). Another observational study by the same group showed a negative correlation between sLF and cortical Aβ load and middle temporal cortex thickness (Reseco, 2021). To date, one clinical trial with sLF in 50 AD patients has been conducted, with results showing improvements in serum Aβ42, ptau181, and inflammatory markers IL-6 and IL-10 after daily intranasal administration of Lf for three months (Mohamed, 2019). While these studies describe promising results, Gleerup et al found no relationship between Lf and AD biomarkers in CSF (Gleerup, 2021). Potential reasons for conflicting results include indiscriminate participant selection, exclusion criteria, and fasting vs non-fasting sample collection.

We investigated the potential relationship between sLF and a comprehensive battery of cognitive tests in a racially diverse, cognitively normal cohort at high risk for AD. Our hypothesis was that lower levels of sLF would correlate to worse cognitive performance, particularly in the domains of memory and visuospatial cognition.

## Materials and Methods

This pilot project was a sub-study of a larger NIH/NIA-funded two-year observational study among cognitively normal, high risk middle-aged adults (PI: Wharton R01AG066203). Recruitment and cognitive testing were conducted by staff members of the Wharton Laboratory as part of the larger study, while sLF collection, processing and analysis were conducted independently (PI: Hammerschlag). Data presented here were collected between October 10^th^ 2022 and March 24^th^ 2023 and all procedures were approved by the Emory Institutional Board (IRB).

Participants taking part in the larger study were invited to take part in this optional salivary collection sub study. We enrolled 17 middle to older-aged (age = 60.29 ± 9.7 years) cognitively normal participants with a biological parent with either autopsy-confirmed or probable AD as defined by NINCDSADRDA criteria (McKhann et al., 1984) and medical records when available. No participant refused the optional salivary collection.

Exclusion criteria included active mouth lesions, a history of dry mouth, contraindication for study procedures, residence in a skilled nursing facility, significant neurological disease including MCI/AD, stroke, history of significant head trauma, untreated Major Depression within 2 years of study enrollment, history of alcohol or substance abuse, current participation in a study with an investigational agent, pregnancy, and unwillingness to fast.

Participants were asked to fast and hold back from other oral stimulation (i.e., smoke, chew gum) for at least 8 hours prior to saliva collection in accordance with previous studies (Bartolome, 2021)(Reseco, 2021), however not all studies, including the above mentioned clinical study reporting no relationship between sLF and cognition (Gleerup, 2021). Saliva samples were collected between 8:00–11:00 AM through the passive drooling technique (Butts, 2022)(Rodriguez, 2023).1-2 mL of unstimulated whole saliva was collected in Falcon™ 15mL Conical Centrifuge Tubes (Corning, Massachusetts, United States). Samples were immediately transported in a cooler with ice packs and centrifuged at 1000 *rpm* for 5 min at 4 °C within 20 minutes of sample collection. Supernatant saliva was then aliquoted into 1.5 ml polypropylene tubes, treated with a protease inhibitor cocktail (cOmplete Ultra Tablets mini, Roche, Basel, Switzerland), and stored at − 81 °C until further analysis in a single batch. Quantification of sLF concentration was performed with a commercially available Human Lactoferrin ELISA Kit (Abcam, ab200015, Cambridge, England), according to the manufacturer’s instructions and in accordance with established protocols (Carro, 2017). All samples were diluted 1:10,000 into sample diluent, after which we added 50 μL of the diluted samples or a standard to the designated wells. All samples were tested in duplicate and the average of the two measurements was used for data analyses. Intra-assay CV values used for analysis differed by an average of two-hundredths of a decimal.

A cognitive battery lasting approximately 90 minutes was administered as a part of the larger parent study, in person, by a trained psychometrician. Cognitive tests were selected based on our and others’ studies in cognitively normal cohorts of individuals at high risk for AD (Kumar, 2020)(Krueger, 2022). Cognitive domains included executive function, memory, visuospatial tasks, attention, and verbal fluency. Testing includes the Forwards and Backwards Digit Span Memory Test (Wechsler, 1981), Mental Rotation Test (Vandenberg & Kuse, 1978), Buschke Memory Test (Buschke, 1973), Consortium to Establish a Registry for Alzheimer’s Disease (Fillenbaum, 1986), Trail Making Test (Bowie & Harvey, 2006), Animal Fluency (Rosen, 1980), Rey–Osterrieth Complex Figure (Rey, A., & Osterrieth, 1941), Judgment of Line Orientation (Benton, 1983), Digit Symbol Substitution Test (Wechsler, 1944), and the Montreal Cognitive Assessment (MoCA) (Nasreddine et al., 2005). In accordance with previous studies, the Forwards Digit Span Memory Test was used as a measure of memory and the Mental Rotation Test as a measure of visuospatial cognition (Richardson, 2007)(Shepard, 1971).

Demographic variables were summarized using descriptive statistics. Multiple bivariate Pearson’s correlations were performed to investigate potential linear relationships between sLF and individual cognitive tests. To control for confounding factors, demographic variables, including age, gender, race, and years of education were adjusted for in all correlations that showed significance. P values were set at 0.05. No correlation lost significance level after adjustment. All statistical analyses were performed by SPSS® ver. 29.0.0.0 (IBM, Armonk, New York, USA).

## Results

|  | **Total Sample**<br>**N = 14** | **Black American**<br>**N = 7** | **Non-Hispanic White**<br>**N = 7** |
| --- | --- | --- | --- |
| **Age** | 60.29 ± 9.7 | 58.7 ± 7.9 | 61.22 ± 11.63 |
| **Sex Assigned at Birth (m/f)** | 4/10 | 2/5 | 2/5 |
| **Education Years** | 16.3 ± 2.2 | 15.7 ± 1.9 | 16.7 ± 2.5 |
| **Salivary LF (ug/ml)** | 18.7 ± 10.7 | 18.2 ± 9.1 | 19.2 ± 12.3 |
| **Cognitive Battery** |  |  |  |
| **Digit Span Memory Test** | 10.5 ± 1.9 | 10.9 ± 2.0 | 10.2 ± 1.9 |
| **Mental Rotation Test** | 17.0 ± 6.1 | 14.4 ± 6.6 | 19.0 ± 5.3 |
| **BSRT delayed recall** | 5.4 ± 2.9 | 5.71 ± 2.5 | 5.2 ± 3.3 |
| **CERAD delayed recall** | 5.2 ± 3.0 | 5.1 ± 2.2 | 5.3 ± 3.6 |
| **TMT-B** | 91.6 ± 53.0 | 81.3 ± 20.5 | 99.6 ± 69.1 |
| **Animal Fluency** | 23.9 ± 6.0 | 21.9 ± 5.0 | 25.6 ± 6.4 |
| **ROCF** | 30.5 ± 5.8 | 28.2 ± 8.2 | 32.3 ± 2.3 |
| **JoLO** | 23.1 ± 4.7 | 21.7 ± 4.0 | 24.2 ± 5.0 |
| **Digit Symbol Substitution** | 59.2 ± 17.2 | 59.3 ± 6.8 | 59.1 ± 22.3 |
| **MoCA** | 26.9 ± 4.2 | 26.71 ± 2.4 | 27.0 ± 5.4 |

*Results are expressed as mean ± standard deviation, unless otherwise stated. Abbreviations: N number, F female; M male, TMT-B Trails Making Test Form B, LF Lactoferrin, CERAD Consortium to Establish a Registry for Alzheimer’s Disease, MoCA Montreal Cognitive Assessment, JoLO Judgment of Line Orientation, ROCF Rey–Osterrieth Complex Figure, BSRT Buschke Selective Reminding Test*.

We analyzed saliva samples and cognitive test data from 14 subjects. **Table 1** shows sample characteristics. Analyzed data consisted of cognitively normal middle to older-aged subjects at risk for AD. Almost 50% of all subjects enrolled in this pilot study are BAs and we analyzed race as a potential modifying variable for general sLF levels.

Age, sex assigned at birth, and years of education were not significantly different between BA and NHWs. Those same variables were used as controls for correlation analyses of sLF and memory.

We found a significant correlation between sLF and the Digit Span Memory Test (DSMT) scores in our cohort’s participants (Pearson’s = 0.607; *P* = 0.013; Fig. 1). The level of significance measured stands after adjusting for age, sex, race, and years of education (*P* = 0.016). The DSMT is used as an assessment of ability to momentarily store and handle information (Richardson, 2007). We suggest that the subjects who had higher levels of lactoferrin in saliva tend to have better memory. This indicates a possible protective effect of lactoferrin toward a cognitive area typically affected by Alzheimer’s Disease.

**Fig 1.**
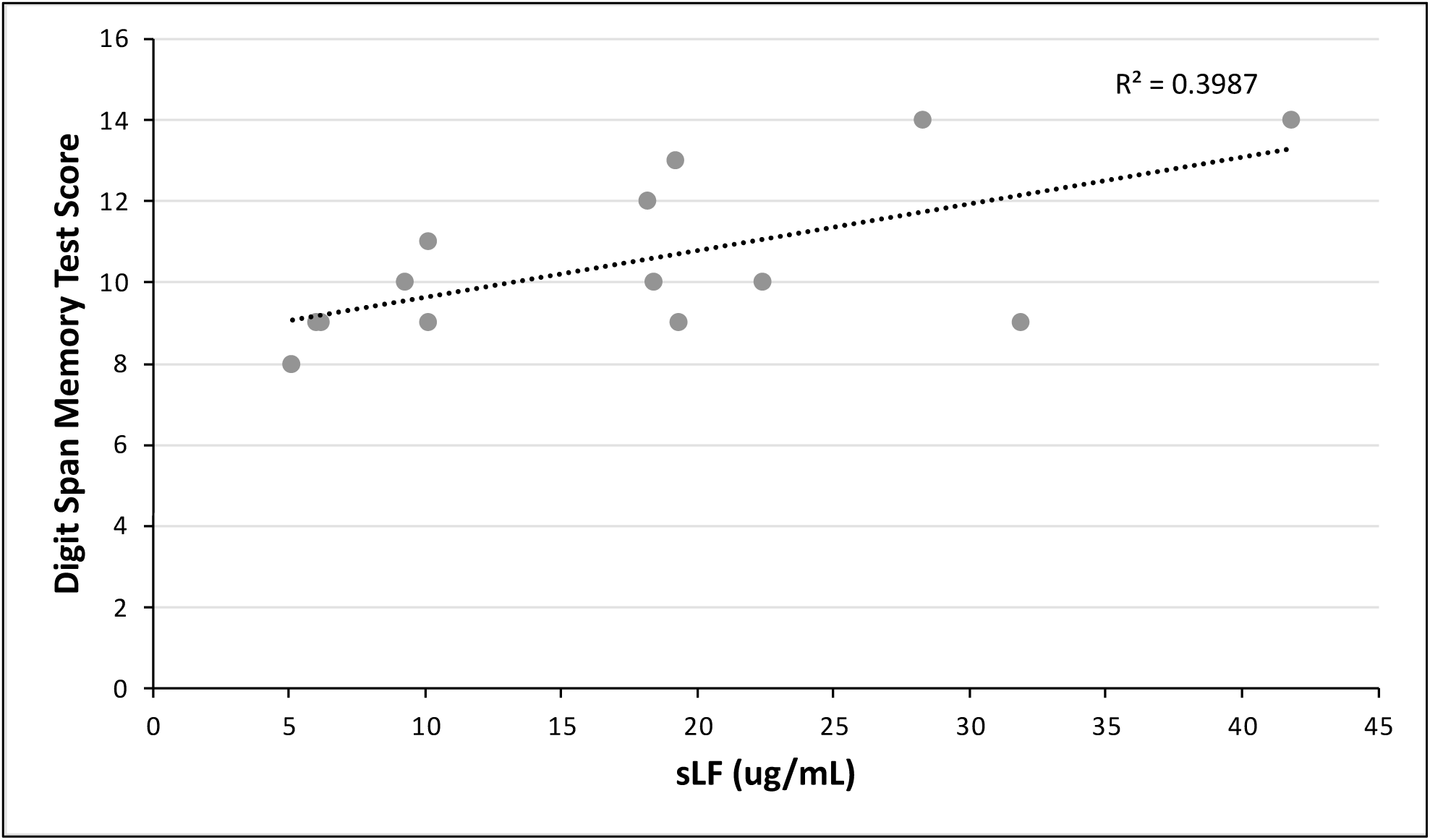
Digit Span Memory Test scores as a function of salivary lactoferrin concentration. We performed a bivariate Pearson’s correlation, and found a positive relationship (P = 0.013).

We found a correlation that is approaching significance between sLF and the Mental Rotation Test scores in our cohort’s participants (Pearson’s = 0.342; *P* = 0.194; Fig. 2). The MRT is used as an assessment of the ability to imagine spatial transformations of objects (Hegarty, 2018). We suggest that subjects who have lower levels of lactoferrin in saliva could have worse spatial cognition. Our small sample size might be restraining us from establishing a statistically significant correlation. It is also worth noting that no other cognitive realm besides memory and spatial cognition had significant or close to significant correlation values.

**Fig 2.**
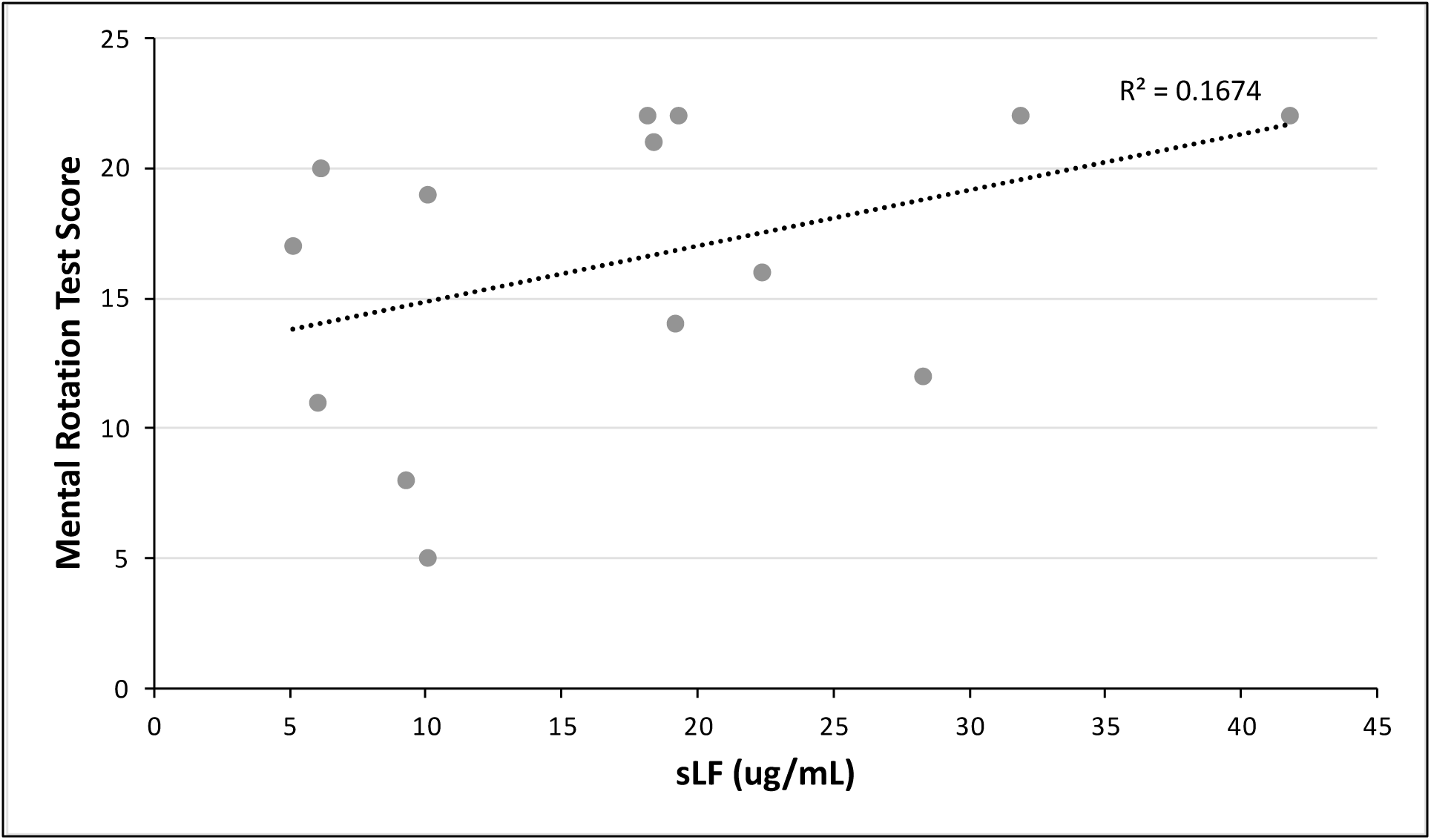
Mental Rotation Test scores as a function of salivary lactoferrin concentration. We performed a bivariate Pearson’s correlation, and a found possible relationship that is approaching significance (P = 0.194).

**Fig. 3.**
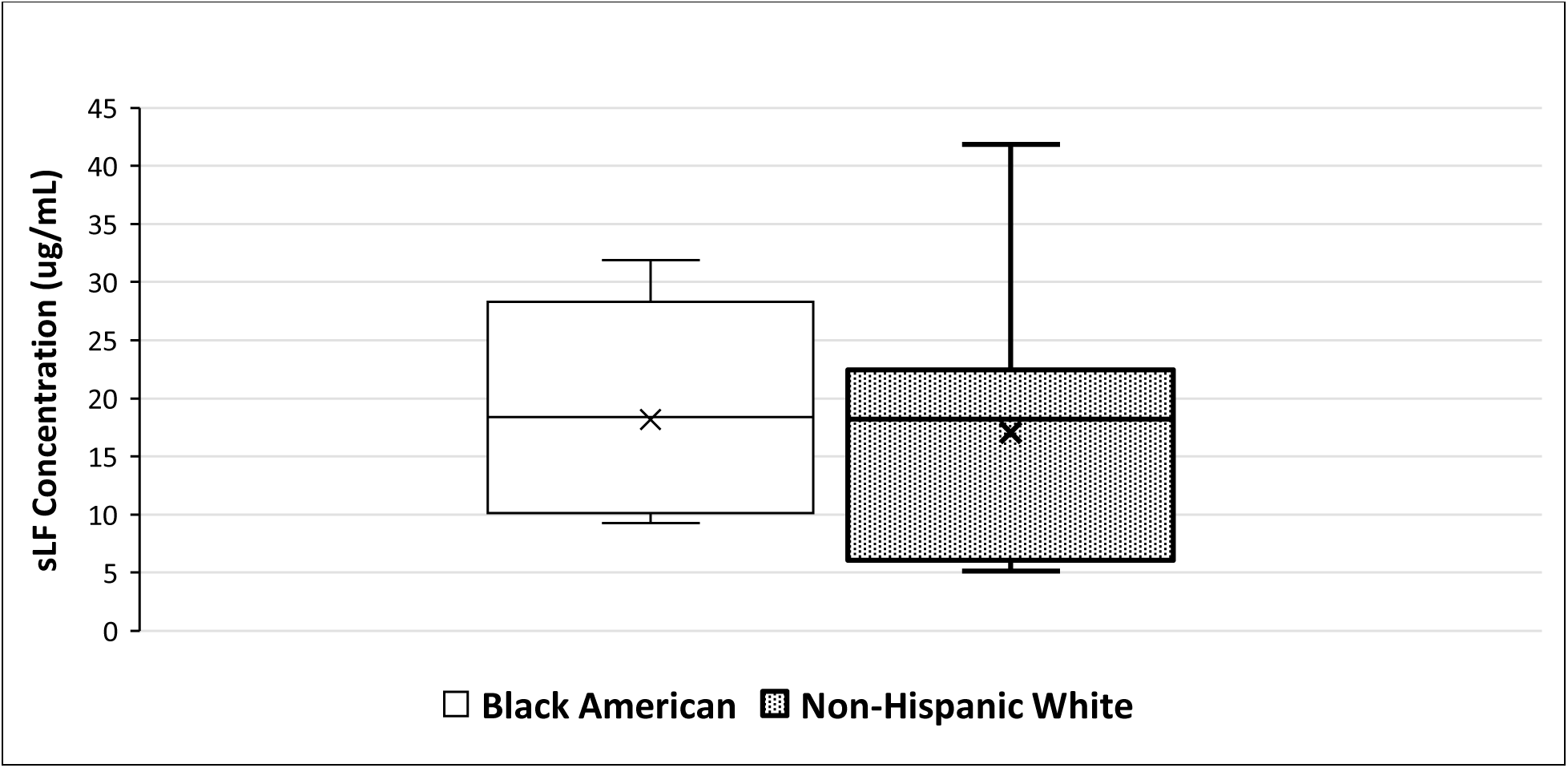
Average salivary lactoferrin levels in Black American and non-Hispanic white participants. There appears to be no significant difference between both groups.

We found no significant difference in average sLF concentration between BAs and NHWs. While each group is composed of only 7 subjects, the average difference is only of ∼5%, as BAs had an average sLF concentration of 18.2 μg/mL and NHWs an average of 19.2 μg/mL.

## Discussion

Conflicting results about the potential relationship between sLf and AD pathology and clinical symptomatology have been reported (Gonzalez-Sanchez, 2020)(Gleerup, 2021). The purpose of this pilot project was to assess the potential relationship between sLf and cognition in a diverse cohort of cognitively normal individuals at risk for AD by virtue of parental history. Secondarily, we explored whether sLf levels differed by race.

Despite our small sample size, we observed a correlation between sLF and memory and visuospatial ability. Cognitive tests were thoughtfully selected based on our prior research showing that they are able to detect subtle cognitive decline in a middle age, high risk cohort (Kumar, 2020)(Nutaitis, 2019). Memory concerns and decline in visuospatial ability (i.e. spatial navigation) are signs of cognitive decline and are classic symptoms of AD (Cacciamani, 2021) (Holger, 2013). Moreover, evidence from animal studies and Lf suggests a domain specific relationship between visuospatial ability and Lf (Zheng et al., 2020). This study suggests a similar relationship in humans — one that, to our knowledge, had not been described before. Difficulties in spatial cognition are common in AD patients, so this possible relationship should be studied further (Coughlan, 2018)(Seo, 2021).

These two conclusions raise questions about the rationale behind the contrasting results between previous studies analyzing sLF and AD (Gonzalez-Sanchez, 2020)(Gleerup, 2021). *Spitting image: can saliva biomarkers reflect Alzheimer’s disease?* is an open-peer commentary that explores the conundrum between the two research groups and the variability in sLF concentration between cohorts (Ashton et al., 2021). The authors question the reproducibility of saliva collection, which can be sensitive to external stimuli and be affected by oral hygiene. We propose a possible alternate explanation for the discordance between the two groups: Gleerup et al. measured sLF levels in non-fasted participants and Gonzalez-Sanchez et al. asked participants to fast overnight. Lactoferrin is found in widely available food items such as meat, cheese, and milk (Superti, 2020), so food intake directly before saliva collection could alter baseline concentrations.

We asked participants to fast overnight prior to saliva collection, but two participants reported having eaten the morning of their visit, so they were excluded from this sub-study. Since the participants had already provided written consent, we still processed these samples, and saw that their sLf levels were 4 standard deviations higher than the average concentration measured in participants included in the sub-study. These post-hoc analyses suggest the importance of fasting before sLf collection. Controlled studies investigating the interaction between fasting and sLF levels are needed to confirm this.

A limitation of this pilot study is its small sample size and that the data presented is correlational and not causal. While inferences can be made about the validity of the protein as a biomarker for AD, results should be taken as preliminary and should be replicated in further studies.

To our knowledge, no previous studies investigating sLF in humans in relation to any neurological disease had reported on race as part of their analysis and/or cohort demographics. As such, our conclusions, which should be replicated across even more races, in other bodily fluids, and with a larger sample size, suggest that sLF has potential to be used as a biomarker for risk assessment in adults predisposed to AD regardless of their race.

Our research group will continue collecting saliva samples from E2 subjects, with the aim of furthering the knowledge and statistical power of the conclusions reached in this pilot study. It is imperative to have easily accessible, disease specific biomarkers that can be clinically useful for determining risk, measuring disease progression and diagnosing AD. We believe that the conclusions reached in this pilot study bring us closer to that goal, but more work is needed.

## Data Availability

All data produced in the present study are available upon reasonable request to the authors

